# Overcoming challenges to successful recruitment of individuals at the earliest stages of disease: An example from Parkinson’s disease research

**DOI:** 10.1101/2025.06.17.25329789

**Authors:** Bridget McMahon, Roseanne Dobkin, Maggie Kuhl, Laura Heathers, Lynell Lemon, Jessica Dimos, Ethan Brown, Kerri A. Pierz, Ken Marek

## Abstract

**Background/Aims:** The Parkinson Progression Markers Initiative (PPMI) is an international, longitudinal, observational study that acquires comprehensive clinical, imaging and genetic data, as well as cerebral spinal fluid (CSF) and blood biological samples from individuals in all stages of Parkinson’s disease (PD) to develop biomarkers that accelerate therapeutic development. The effort to enroll a large population of prodromal individuals (participants with PD biomarkers and/or early symptoms) presents an opportunity to share insights for overcoming specific challenges unique to enrollment prior to the onset of typical symptoms.

**Methods:** Paid and earned media, referral cohorts, and individual and community ambassadors were leveraged as recruitment strategies to attract and enroll a broad study population. The success of each strategy was measured as conversion to (i) informed consent, (ii) smell test completion, and (iii) identification of hyposmic individuals. Participants completed an eConsent and questionnaire online, followed by remote smell test screening for Parkinson’s risk. Participants with hyposmia were invited to complete dopamine transporter (DAT) imaging. Those who continued to meet eligibility criteria were invited to enroll in the PPMI clinical study, for comprehensive longitudinal follow-up.

**Results:** Over 1.2 million visits to the smell test website were recorded and 79,837 participants consented to remote hyposmia screening. A smell test was sent to 57,754 participants and completed by 38,419; 6,649 were identified to be hyposmic (as defined by PPMI study cutoffs). DAT imaging was completed by 1,720, with 1,120 identified as eligible for longitudinal follow-up. The greatest number of participants who consented to smell testing came from paid media and social media ads, referral networks (commercial and study partners), and the MJFF community. Overall, 14.4% of participants who completed smell testing were confirmed to be hyposmic. The highest rates of conversion from completed UPSIT to confirmed case of hyposmia occurred with promoted online search results, earned press coverage, and radio ads.

**Conclusion:** PPMI provides a useful example of overcoming challenges to enrollment of participants with biomarkers and/or early symptoms (but without typical symptoms) of disease. Important lessons from these data for this study team, and other researchers alike, are to be agile in recruitment strategies, to utilize effective participant engagement approaches, and to prioritize centralized study teams/resources to offload site burden and streamline participant experience.

## BACKGROUND

Timely and effective participant recruitment in clinical trials is key to study success and under-recruitment for clinical trials slows medical research progress and increases expenses.^1^ Approximately one-half (40% - 60%) of clinical studies fail to meet enrollment targets,^1^ and low accrual rates are the most common single reason trials are prematurely terminated.^2^ Studies enrolling individuals at the earliest stage of disease, often before typical symptoms arise enabling a formal clinical diagnosis, pose additional challenges. Traditional recruitment methods are limited in these not yet clinically diagnosed individuals as they are unlikely to present to medical care. These individuals may not recognize their early symptoms or be aware of their personal risk. Thus, complex and resource-intensive measures are required to screen very large numbers of individuals to identify those who possess biomarkers or clinical characteristics of interest. Additionally, it is crucial — even more so in an undiagnosed population — to provide needed education and address concerns that may otherwise interfere with participation. Operational workflows can be designed to identify and engage potential participants, including the use of online and remote screening tools.

We have developed an enrichment strategy to recruit individuals without typical symptoms in the Parkinson’s Progression Markers Initiative (PPMI; https://www.ppmi-info.org/), sponsored by The Michael J. Fox Foundation for Parkinson’s Research (MJFF). PPMI is an international, longitudinal study that acquires comprehensive clinical, imaging and genetic data, as well as CSF and blood biological samples from individuals in all stages of PD, including participants with biomarkers prior to symptom onset and those with functional impairment.^3^ PPMI makes real-time data available for discovery and validation by others. Participants may also provide information about wellness and disease through an online platform. Insights and scientific advances from PPMI have already had a significant impact on clinical trial design, have transformed our definition of PD, and have accelerated PD therapeutic development (https://www.ppmi-info.org/publications-presentations/publications).

Evidence from longitudinal population and biomarker studies in PD and related disorders like Dementia with Lewy bodies (DLB), has demonstrated that neurodegeneration begins years before typical symptoms are present. Recent studies show that the CSF synuclein seed amplification assay (SAA) detects aggregated synuclein in individuals even before symptoms arise and have led to the integrated biomarker and clinical Neuronal Synuclein Disease – integrated staging system (NSD-ISS), encompassing both PD and DLB.^3, 4^ Further, data strongly suggest that a simple test for olfactory dysfunction may substantially enrich for individuals who are alpha synuclein SAA positive.^5–7^

This paper describes recruitment strategies and operational workflows that support large-scale identification, centralized screening, and enrollment of study participants with biomarkers and/or early symptoms enriching for progression to typical symptoms of PD. We describe the development and operational detail for Smell Test Direct (ST Direct), a simple online platform enabling individuals from the community to participate in a research screening paradigm. In PPMI, centralized risk screening and site referrals have been used to (i) engage a broader and more inclusive population of potential participants, (ii) speed identification of larger volumes of individuals with early-stage disease biology, and (iii) reduce site burden. We believe that these strategies and learnings are of significant value to researchers interested in reaching geographically and demographically diverse participant populations and/or efficiently screening a general population.

## METHODS

### Recruitment Goals

To provide an in-depth understanding of the biology and the progression of PD, it is necessary to enroll a large, diverse cohort, that is characterized both with biomarkers and clinical outcomes, from which specific studies can be conducted with selective subsets of participants. The aims of the PPMI recruitment efforts were to minimize upstream and downstream barriers to engagement on the part of the participants and study sites to enable enrollment of a very large cohort of individuals with biomarker evidence of NSD (n ≈ 2,000). The study goal is to longitudinally follow these individuals to determine their disease progression to typical neurodegenerative phenotypes including PD or DBL.

### Participants

Use limited eligibility criteria to identify the broadest population of eligible participants Recruitment tools for PPMI have evolved to meet the needs to enroll larger numbers of study participants. Recruitment for participants prior to the onset of symptoms launched in November 2020 targeting individuals from the community aged ≥60 years without a clinical diagnosis of PD and at least one of the following: (1) a first-degree family member with PD, (2) a diagnosis of REM sleep behavior disorder (RBD) or self-reported dream enactment, or (3) previous genetic testing that identified a pathogenic PD variant. Individuals were directed to the MJFF website to complete a short screener, and if eligible, directed to participate in remote hyposmia screening since smell loss enriches for risk for PD.^8–11^ In July 2021, the referral workflow was modified to direct people to a new PPMI Online data collection platform; from the same self-reported criteria to enlarge the population to be screened and speed the workflow. Eligible candidates were directed to remote hyposmia screening with the University of Pennsylvania Smell Identification Test (UPSIT).

In June 2022, the screening paradigm evolved further focused on additional evidence that hyposmia markedly enriches for PD risk.^8–11^ All adults (≥60 years old) in the United States (US) without a PD diagnosis were eligible to receive an UPSIT via a new streamlined Smell Test (ST) Direct initiative, which enabled rapid community identification of qualified individuals.

Interested individuals could go to mysmelltest.org to complete a screening questionnaire and consent to remote hyposmia screening. Individuals with hyposmia (defined by a data-driven eligibility cutoff) were referred for further participation in PPMI as below.

### Recruitment Strategies

Robust, multimodal strategy was employed to engage target participants See **Error! Reference source not found**. for overview of recruitment methods and participant flow.

MJFF has led a multifaceted PPMI marketing and recruitment strategy around PD awareness, education, and action. MJFF worked with the PPMI Community Advisory Board, the MJFF Patient Council (both comprising individuals of varied demographic and disease backgrounds), and other community members to review messaging, materials, and tactics. Strategies described below include paid media, referral networks, and individual or community-based ambassadors deployed primarily in the US. While some efforts have also been piloted in Canada and the United Kingdom, only results from the US efforts are detailed below.

Since the launch of Smell Test Direct, campaigns have used distinct vanity URLs (e.g., mysmelltest.org/paper) and QR codes to track performance activity. These data allow for an iterative strategy toward greatest success rate and return on investment.

1. Paid and Earned Media Efforts Paid digital media advertising included promoted search engine results, social media ads, and display ads on websites and in gaming apps. Television ads ran in select markets during airings of shows including “Wheel of Fortune,” “CBS Mornings,” and “Family Feud.” Print advertisements in newspapers, community newsletters and alumni magazines were also used, as were radio advertisements. MJFF engaged press for earned coverage of the study. All placements targeted users and readership with age eligibility and, many, with aligned or enriched interests (e.g., searching sleep aids; reading medical sites or materials for veterans who may be exposed to environmental risk factors); some campaigns deployed in geographies with higher volumes of underrepresented populations in service of PPMI’s aim to build an inclusive study population. Ad messaging is critical to driving participant engagement; ad messaging focused on empowerment and impact (e.g., “He doesn’t have Parkinson’s. But he can help end it.”) and simplicity of action (e.g., “A free scratch-and-sniff test can help researchers learn more about brain health.”). The call-to-action directed interested individuals to websites to learn more or submit their information to determine eligibility (i.e., websites of MJFF, Smell Test Direct, and PPMI Online).
2. Referral Cohorts The Foundation has tapped into existing cohorts of people interested in research participation. This includes its own audience; MJFF sent information about PPMI to its own email list and posted to its Facebook community, comprising people with PD and their loved ones. Its website holds content on disease risk, PPMI, and how to get involved. MJFF also pursued partnerships to promote Smell Test Direct. MJFF contracted companies like Quest Diagnostics, SubjectWell, and ClinicalConnection to email their constituencies of people aged 60 and older without PD who opted into research opportunity communications. Longitudinal studies of aged individuals ( i.e., UCSF Brain Health Registry, Alzheimer’s Prevention Registry, 23andMe) are also partnering to refer participants to PPMI.
3. Individual and Community-based Ambassadors Recruitment included collaboration with community-based organizations and individual ambassadors, especially to engage underrepresented populations. MJFF has directly connected with more than 30 community-based organizations. Study staff participated in online educational and in-person events (e.g., health fairs, support groups) hosted by these organizations. MJFF created a partner promotion toolkit with sample social media posts, email and newsletter text, and graphics and images for partner organizations to message about PPMI on their own channels. PPMI study staff and current volunteers (with and without PD) were asked to spread the word of Smell Test Direct. A consultant was engaged to promote Smell Test Direct in active aging resident communities. Working with leadership at more than 25 senior living communities, the consultant trained residency staff and presented the study opportunity through in-person sessions and resident communications (e.g., emails, print newsletters).

### Study Screening Activities

#### Establish PPMI Account

Participants from ST Direct portal, the MJFF portal, or PPMI online were funneled to an online site hosted by Indiana University (IU) and instructed to generate or login to an existing PPMI account.

#### Review and Sign eConsent

After initial log in, participants were required to review and sign the online informed consent form (ICF). Participants could indicate if they had questions before consenting. This would halt their progress until an IU team member spoke with them. Participants could print a signed copy directly from the portal and receive a signed copy via email.

#### Complete Detailed Questionnaire

Participants completed contact details and demographic information on the portal. They were directed to complete a questionnaire that addressed four key, high-interest PD risk factors such as RBD diagnosis, dream enactment, subjective smell loss, and family history of PD.

#### Perform Remote Hyposmia Screening

An UPSIT, a 40-item multiple choice scratch and sniff test, and detailed instructions on how to complete the UPSIT and enter responses in the online portal were mailed to participants. The portal contained UPSIT data entry pages designed to mirror the paper booklets, including the same color coding of pages, to ease online entry of responses by participants. Participants were required to provide responses for all 40 UPSIT questions and instructed to make a guess if the detected odor was not an available multiple-choice option.

#### Determine Ongoing Eligibility and Introduce DaTscan

All remotely collected participant information was used to evaluate each participant’s eligibility to proceed to additional PPMI research assessments. Participants who continued to meet all criteria including evidence of hyposmia were contacted via phone by the IU study team to discuss the next screening step toward clinical study enrollment. Participants were offered the opportunity to go to a PPMI site for dopamine transporter (DAT) imaging using single photon computerized tomography (SPECT) and isoflurane. The IU study team made multiple attempts via phone and email to reach these participants, tracking contacts in the study database. Calls followed an established discussion guide to ensure consistent communication of essential information and to ensure setting appropriate expectations for before, during and after DAT imaging. Additional screening information specific to DAT imaging was acquired. Instances of unclear eligibility were escalated to the PPMI medical monitor for review. This process allowed for the creation of a centralized and shared reference list of medical conditions and medications that would be exclusionary based on the PPMI medical monitor’s assessment, thus streamlining the DAT imaging screening process for subsequent participants.

#### Refer to In-Person Visit at Study Site

At the end of the DaTscan introductory call, participants agreed to a referral to a study site for DaTscan screening, declined referral to a study site, or remained undecided. A follow-up call was scheduled for undecided participants. Participants who declined the DaTscan screening were encouraged to participate in other research opportunities within the PPMI program. For individuals who agreed to DaTscan screening, the IU team collected travel preferences and determined the study site that both minimized participant travel burden and considered site capacity. Identified participants were commonly located far distances from PPMI clinical sites and required travel coordination to complete their study visits. Participants could also designate a travel companion to accompany them on study visits. All costs related to the study visits were covered by PPMI, with an expense policy provided to participants in advance. IU referrals were posted to a secured platform shared with site teams, with each team having restricted visibility to only their assigned participants. This secured platform was used for direct communication between the IU team and site teams about individual referrals.

PPMI site teams were responsible for scheduling visits and submitting visit details to the IU team. The IU team partnered with a travel vendor to organize required travel and to generate individual itineraries for participant visits. The IU team monitored visit rescheduling and cancellations, updated itineraries, and organized reimbursements for participant-incurred expenses. The travel team was available outside of normal business hours to support participants during unexpected travel disruptions, such as flight delays.

Determine Clinical Eligibility for Enrollment and Longitudinal Follow-up: Following completion of DAT imaging, images were quality controlled and quantitatively analyzed centrally by the PPMI imaging core. Participants were eligible to be fully enrolled in PPMI based in quantitative DAT striatal specific binding cutoffs adjusted for age and sex. The IU study team scheduled follow-up phone calls with participants to explain the comprehensive longitudinal follow-up required for PPMI and to assess study participation interest.^12^ If they agreed, the IU study team reconfirmed travel and other personal preferences to organize site visit logistics. Ineligible participants were contacted and thanked for their participation. Other opportunities for continued participation in PPMI were offered.

#### Participant Communications

To help address participant concerns or anxiety, standardized talking points and educational resources were developed in partnership with study participants, IU, and site study teams. Central development of communication materials ensured consistency in tone and messaging of study details and impact (e.g., talking points, flyers, brochures, welcome packets, slides for investigators). At each step in the process, participants had access to trained staff to explain next steps.

### Site Selection and Management of Workload

From among the 31 PPMI sites in the US, 12 referral sites were contracted to perform the initial DAT imaging screenings. Sites with long-term experience working on PPMI, engaged nuclear medicine teams, and ability to schedule and conduct a high number of scans were chosen to support the high-throughput DaTscan screenings required. Typically, one dedicated site study coordinator managed these screening visits, including scheduling, consenting, and supporting participants through the visit. While participants were screened using DAT imaging at high throughput referral sites, PPMI participants could undergo their baseline assessment at any of the 31 US PPMI sites.

The PPMI Site Management Core (SMC) worked with sites to help increase capacity for DaTscan screenings and/or for accepting baseline-eligible participants to ensure all participants would be seen at sites within 4-6 weeks from referral. SMC utilized a shared spreadsheet with the IU team to communicate current site information and track monthly capacity for DaTscan and baseline visits. Regular communication between IU and SMC teams facilitated successful management of site capacity and participant referrals.

Monthly metrics of site progress were sent to all referral sites. SMC met regularly with referral sites to ensure referrals were managed efficiently and to minimize operational obstacles. As the number of referrals expanded and more US sites received baseline-eligible participants, both IU and the SMC maintained regular contact and oversight of the referrals.

## RESULTS

### Results of recruitment strategies

As of the end of Q1 2024, 100,602 total individuals have created PPMI accounts for remote smell testing, referred collectively through ST Direct, the MJFF screener and PPMI Online, resulting in 79,837 who completed consent for remote smell testing. This consented population had a mean age of 68 years (range: 60-100 y). This population was majority female (77.4%), White race (73.8%), and not Hispanic or Latino (71.9%). See Error! Reference source not found. for additional demographic information.

Additionally, 62,149 completed the online questionnaire about PD risk, and 57,754 were mailed the UPSIT to perform remote hyposmia screening (of which, 67% were completed and returned). Of those who completed UPSITs (n=38,419), 12% (6,649) were determined to have hyposmia; n=2,833 of whom agreed to further screening with a DaTscan. Over 60% (n=1,720) of those eligible have thus far completed the DaTscan, and 65% of those (n=1,120) determined to be eligible to continue into in-clinic long-term follow-up. The total yield is 11% of all individuals identified as hyposmic who enrolled in the longitudinal follow-up phase of the study. Overall outcomes of the enrollment activities and phase screening process are summarized in the funnel plot found in Error! Reference source not found..

Since its launch in June 2022, the mysmelltest.org site (Smell Test Direct) has seen more than 1 million page hits. MJFF closely monitored the performance of varied recruitment campaigns to allow changing strategies over time (see also Table 1 and Table 2). The results below on the effectiveness of the recruitment strategies focus only on Smell Test Direct recruitment; earlier recruitment strategies have been replaced by ST Direct. Overall, paid and earned media strategies contributed the greatest proportion of total ST Direct consented participants (50% of total), completed UPSITs (44% of total), and identified hyposmic individuals (50% of total) (see Figure 2). Additional details of each strategic approach are further described below.

1. Paid and Earned Media Efforts Paid and earned media efforts have led to a high volume of consents for remote smell testing (n=35,912). As of the end of Q1 2024, 42% (n=14,989) of these ST Direct consented participants completed the remote hyposmia screening using the UPSIT, resulting in the identification of 2,481 hyposmic participants who remained eligible to proceed to subsequent steps. To start, MJFF tracked paid media as an amalgam of efforts. Once more nuanced tracking was employed, reports showed Facebook ads contributed the highest number of consents for smell testing compared to other social media channels (e.g., Twitter, Reddit). The greatest conversion to consents was found via direct mail strategies; although only 1381 page visits resulted from direct mail efforts, 28% (383) were completed the online consent (followed by print newspaper ads, from which 21% of site visitors completed the consent for smell testing). Other channels such as native and display ads were successful in driving page visits, but those visits converted to consents at a low rate (∼1%). Television (within the Paid Media General Campaign category), radio and print advertisements drove fewer consents but may have contributed to general awareness of the study.
2. Referral Cohorts Recruitment through referral cohorts has led to a high volume of ST Direct participants consented for smell testing (n=30,164; 42% of the 71,953 consented from ST Direct). As of the end of Q1 2024, 53% (n=15,994) of consented participants from these referral cohorts completed the UPSIT, resulting in the identification of 1,967 hyposmic participants who remained eligible to proceed to subsequent steps in the protocol. Through these referral cohorts, 14,191 and 15,973 participants have consented to smell testing from recruitment through the MJFF community and the third-party efforts (i.e., commercial companies, partner studies), respectively.
3. Ambassadors Recruitment through ambassadors led to 2,108 consents (3% of total participants consented through ST Direct). As of the end of Q1 2024, 54% (1,136) of consented participants from these recruitment strategies completed the remote hyposmia screening using the UPSIT, resulting in the identification of 144 hyposmic participants who remained eligible to proceed to subsequent steps in the protocol. Among those from ST Direct who completed remote smell testing (n=34,174), 14% (n=4,938) were identified as hyposmic (Error! Reference source not found.). The recruitment methods that yielded the greatest proportion of hypsomic participants from the total consented participants from ST Direct included earned press coverage, radio ads, in-person MJFF events, and PPMI staff ambassadors (each with at least 10% conversion from consent for smell testing to confirmed hyposmia). Referrals through third-party commercial providers and from partner studies had among the lowest rates of conversion from consent to hyposmia (≤6%) of all methods explored. (Data not shown)

**Figure 1.**
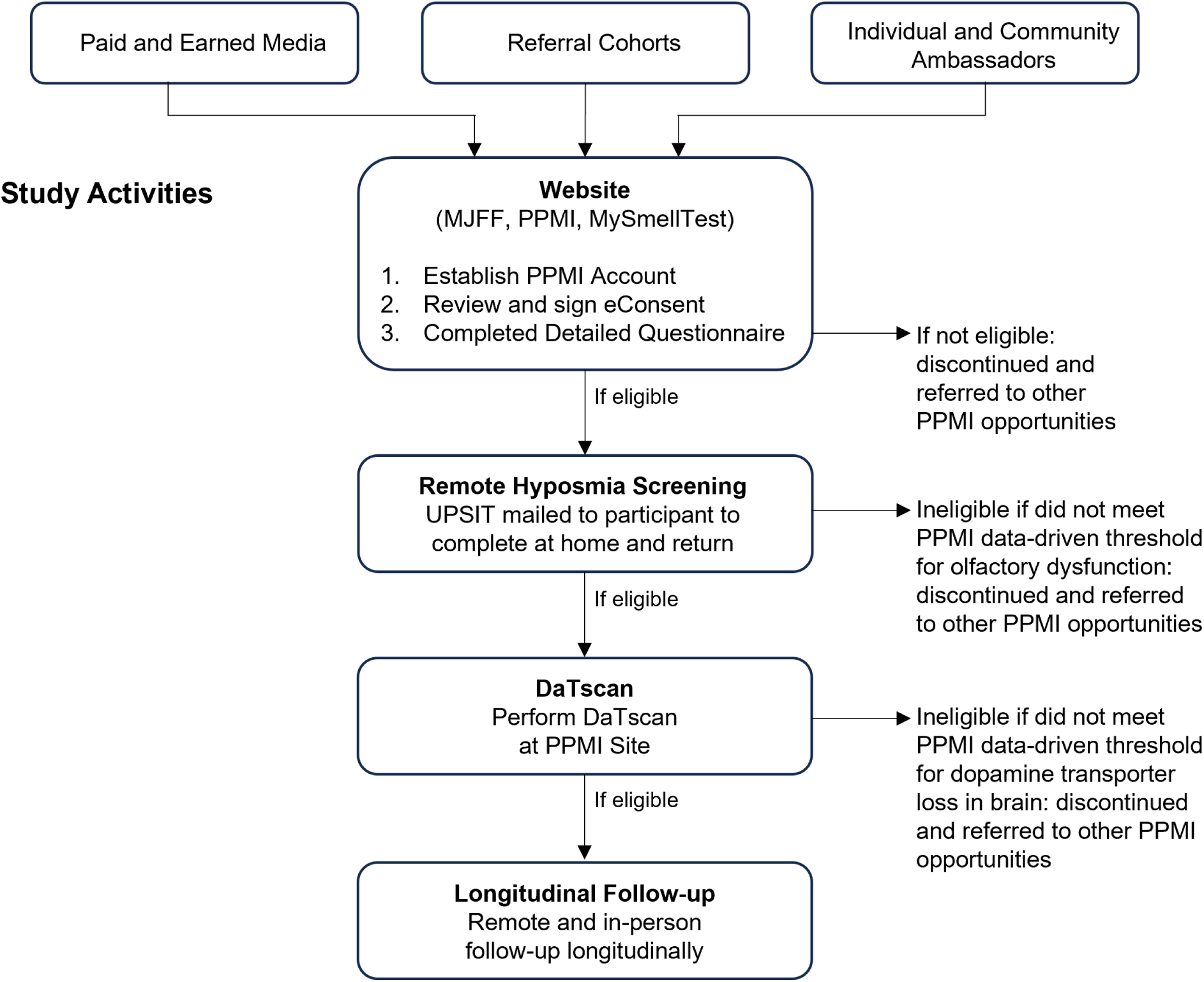
Overview of recruitment and engagement methods. Multi-modal recruitment included paid and earned media, referral cohorts, and individual/community ambassadors, which all funneled into a website where potential study participants could complete background information and review/sign the electronic informed consent. The subsequent eligibility steps and process are depicted in the figure culminating in the selection of study participants with confirmed olfactory and dopamine deficits.

**Figure 2.**
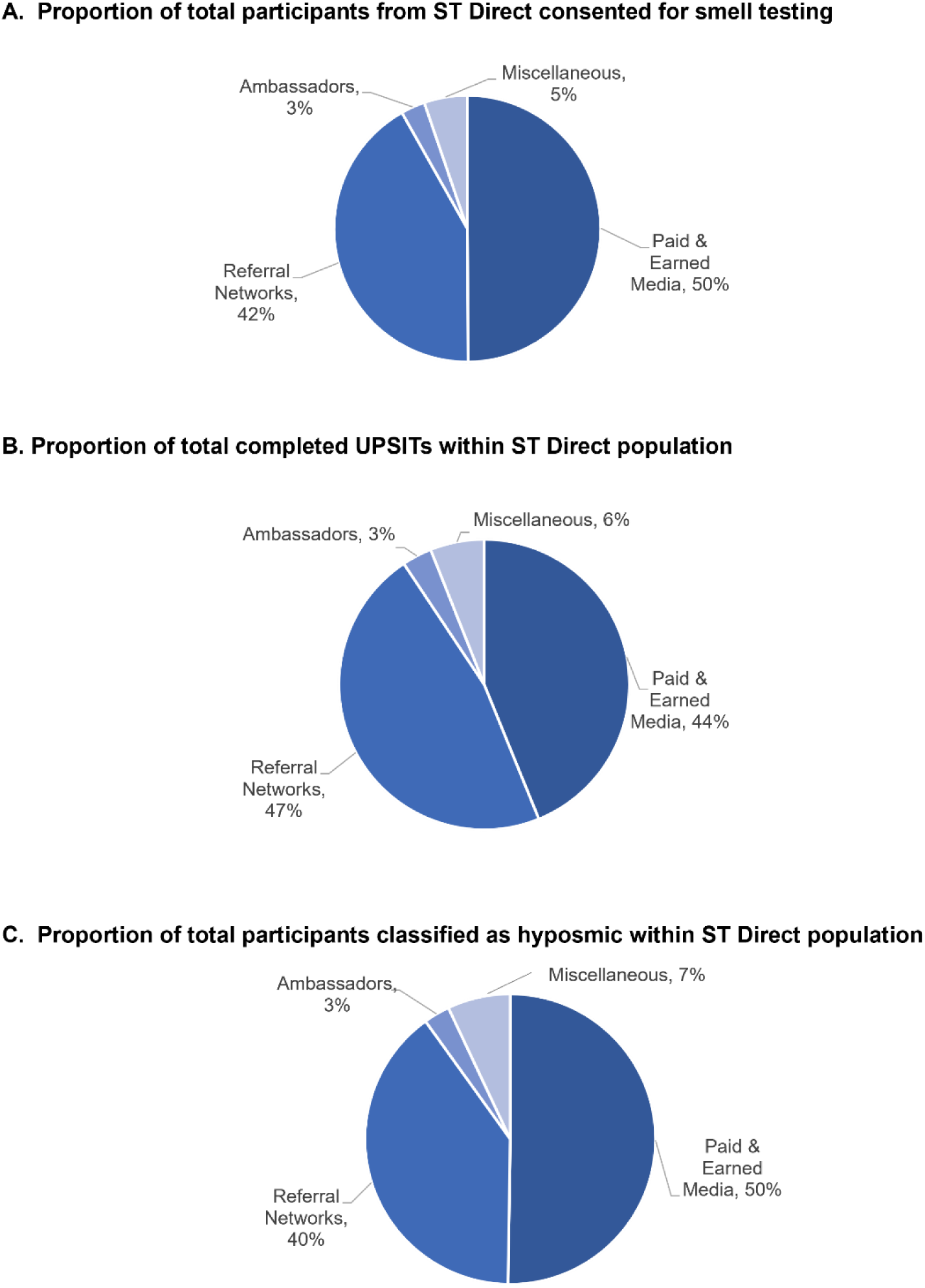
Contribution of each strategy to recruitment efforts within the ST Direct population. The proportion of total ST Direct participants who consented to smell testing by each recruitment strategy (A), proportion of total ST Direct participants who completed the UPSIT by each recruitment strategy (B), and proportion of total ST Direct participants classified as hyposmic (C) are depicted in the pie charts shown.

**Figure 3.**
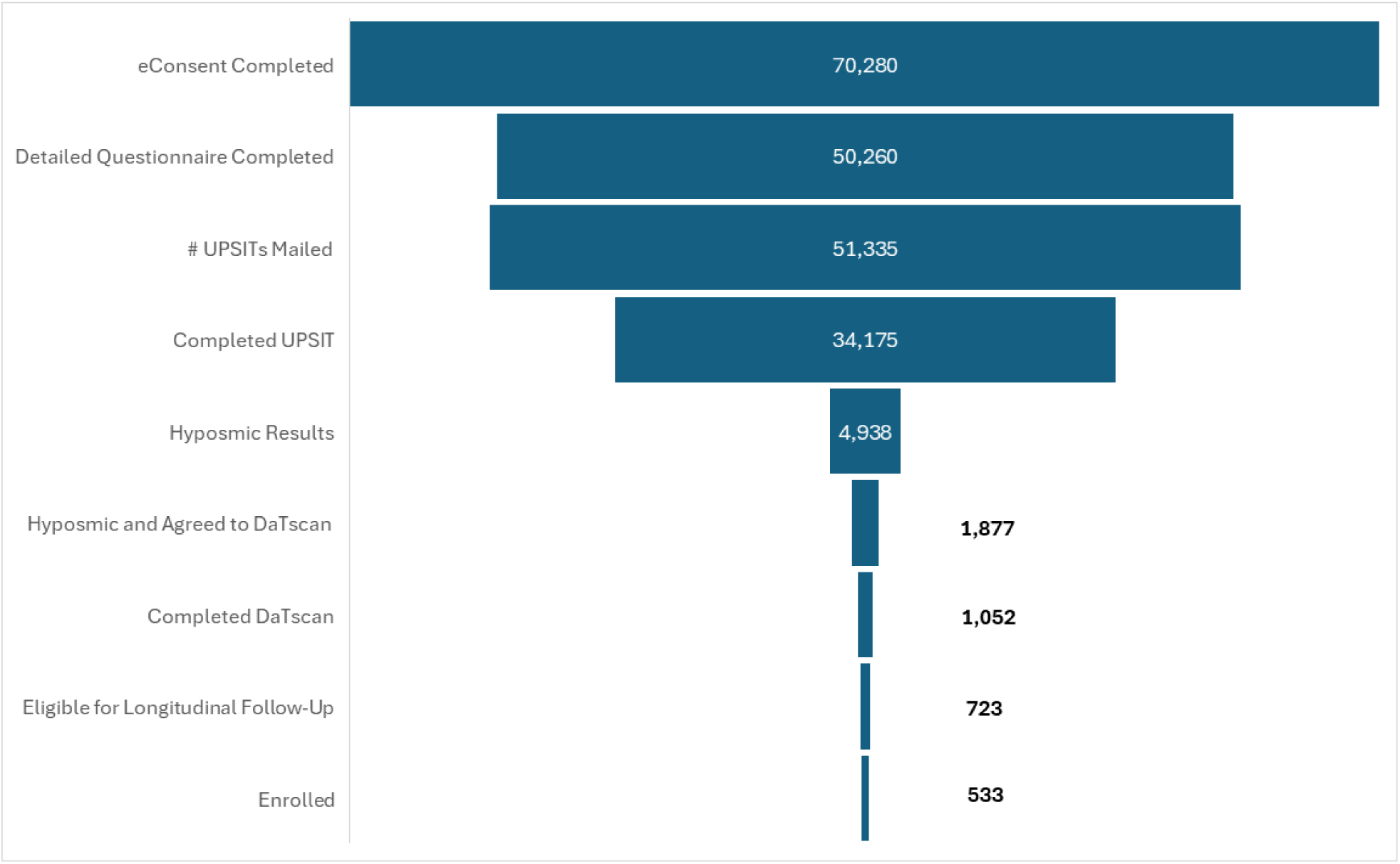
Participant flow through study recruitment and enrollment phases for total population. Attrition of study participants through each phase of the recruitment and enrollment process is depicted in the figure, resulting in 533 participants enrolled in longitudinal follow-up.

**Table 1.** Demographics of study population who completed electronic consent for remote smell testing.

| **Consented population** |  |
| --- | --- |
| N | 79,837 |
| **Age at consent, y** |  |
| Mean | 68 |
| Range | 60-100 |
| **Sex, n (%)** |  |
| Male | 18,071 (22.6) |
| Female | 61,766 (77.4) |
| **Race, n (%)** |  |
| American Indian or Alaska Native | 187 (0.2) |
| Asian | 612 (0.8) |
| Black or African American | 796 (1.0) |
| Native Hawaiian or Other Pacific Islander | 46 (0.1) |
| White | 58,920 (73.8) |
| More than one race indicated | 582 (0.7) |
| Not available | 18,694 (23.4) |
| **Ethnicity** |  |
| Hispanic or Latino | 1,761 (2.2) |
| Not Hispanic or Latino | 57,403 (71.9) |
| Not available | 20,673 (25.9) |

**Table 2.** Outcomes of ST Direct recruitment strategies to enroll participants at the earliest stage of disease.

| **Campaign** | **Page Visits**<br>**(n)** | **Signed**<br>**Consent**<br>**(n)** | **Completed UPSIT**<br>**(n)** | **Classified as**<br>**Hyposmic**<br>**(n)** |
| --- | --- | --- | --- | --- |
| **Paid & Earned Media** |  |  |  |  |
| Paid Media General Campaign | 123,019 | 19,407 | 8,002 | 1,480 |
| Social Media Ads | 482,937 | 13,304 | 5,481 | 754 |
| Promoted Online Search Results | 103,922 | 1,000 | 409 | 94 |
| Print Newspaper Ads | 3,720 | 794 | 426 | 53 |
| Website Display Ads | 107,868 | 754 | 318 | 36 |
| Direct Mail | 1,381 | 383 | 193 | 30 |
| Earned Press Coverage | 1,274 | 246 | 148 | 33 |
| Point of Care Ads | 75 | 14 | 7 | 0 |
| Radio Ads | 131 | 10 | 5 | 1 |
| **Category Totals** | **824,327** | **35,912** | **14,989** | **2,481** |
| **Referral Networks** |  |  |  |  |
| *Third Parties* |  |  |  |  |
| Commercial Companies | 61,497 | 9,995 | 4,994 | 487 |
| Partner Studies | 9,384 | 4,279 | 2,741 | 246 |
| Senior Living | 4,268 | 1,217 | 609 | 104 |
| Nonprofit Partner Contracts | 1,682 | 482 | 301 | 44 |
| *MJFF Community* |  |  |  |  |
| MJFF Email List | 55,162 | 7,115 | 3,510 | 503 |
| MJFF Website Content | 37,932 | 6,623 | 3,612 | 545 |
| MJFF Facebook Community | 3,414 | 373 | 181 | 30 |
| In-person MJFF Events | 537 | 80 | 46 | 8 |
| **Category Totals** | **173,876** | **30,164** | **15,994** | **1,967** |
| **Ambassadors** |  |  |  |  |
| Partners Toolkit | 7,430 | 1,161 | 588 | 70 |
| PPMI Site | 6,966 | 447 | 240 | 35 |
| PPMI Participants | 2,350 | 302 | 196 | 19 |
| PPMI Staff Member | 1,621 | 198 | 112 | 20 |
| **Category Totals** | **18,367** | **2,108** | **1,136** | **144** |
| **Miscellaneous** |  |  |  |  |
| General URL entry | 31,367 | 3,701 | 2,027 | 333 |
| Unknown | 417 | 68 | 28 | 13 |
| **Category Totals** | **31,784** | **3,769** | **2,055** | **346** |
| **GRAND TOTAL** | **104,8354** | **71,953** | **34,174** | **4,938** |

**Table 3.** Success rate of each ST Direct recruitment strategy to have remote smell testing completed and successfully identify hyposmia among the study participants.

| **Campaign** | **Completed UPSIT (n)** | **Classified as Hyposmic (n)** | **Conversion from Completed UPSIT to Classified Hyposmia** | **% of Total Completed UPSITs\*** | **% of Total Classified as Hyposmic\*** |
| --- | --- | --- | --- | --- | --- |
| **Paid & Earned Media** |  |  |  |  |  |
| Paid Media General Campaign | 8,002 | 1,480 | 18% | 23% | 30% |
| Social Media Ads | 5,481 | 754 | 14% | 16% | 15% |
| Promoted Online Search Results | 409 | 94 | 23% | 1% | 2% |
| Print Newspaper Ads | 426 | 53 | 12% | 1% | 1% |
| Website Display Ads | 318 | 36 | 11% | 1% | 1% |
| Direct Mail | 193 | 30 | 16% | 1% | 1% |
| Earned Press Coverage | 148 | 33 | 22% | 0% | 1% |
| Point of Care Ads | 7 | 0 | 0% | 0% | 0% |
| Radio Ads | 5 | 1 | 20% | 0% | 0% |
| **Referral Networks** |  |  |  |  |  |
| *Third Parties* |  |  |  |  |  |
| Commercial Companies | 4,994 | 487 | 10% | 15% | 10% |
| Partner Studies | 2,741 | 246 | 9% | 8% | 5% |
| Senior Living | 609 | 104 | 17% | 2% | 2% |
| Nonprofit Partner Contracts | 301 | 44 | 15% | 1% | 1% |
| *MJFF Community* |  |  |  |  |  |
| MJFF Email List | 3,510 | 503 | 14% | 10% | 10% |
| MJFF Website Content | 3,612 | 545 | 15% | 11% | 11% |
| MJFF Facebook Community | 181 | 30 | 17% | 1% | 1% |
| In-person MJFF Events | 46 | 8 | 17% | 0% | 0% |
| **Ambassadors** |  |  |  |  |  |
| Partners Toolkit | 588 | 70 | 12% | 2% | 1% |
| PPMI Site | 240 | 35 | 15% | 1% | 1% |
| PPMI Participants | 196 | 19 | 10% | 1% | 0% |
| PPMI Staff Member | 112 | 20 | 18% | 0% | 0% |
| **Miscellaneous** |  |  |  |  |  |
| General URL Entry | 2,027 | 333 | 16% | 6% | 7% |
| Unknown | 28 | 13 | 46% | 0% | 0% |
| **GRAND TOTAL** | **34,174** | **4,896** | **14%** | **100%** | **100%** |

\*Within the population recruited from ST Direct only.

## CONCLUSIONS

PPMI serves as a useful case of real-world experience utilizing broad-reaching efforts to overcome the challenges frequently associated with recruiting study participants prior to the onset of symptoms. We highlight the strategy to deploy ST Direct using a simple, widely available, relatively inexpensive, at-home test to screen for PPMI participants. The degree of attrition through the steps in the process reinforces the need to reach and screen a great number of individuals to produce a sufficiently large target cohort. We have extensively investigated recruitment strategies in PPMI to provide guidance for other researchers who encounter similar challenges in the study of early disease.

To date, the greatest yield of consented participants numerically was through paid social media ads, third-party referral partners, and the MJFF community (Error! Reference source not found.). Correspondingly, the largest proportion of total completed smell tests and identification of individuals with hyposmia were from the same recruitment strategies (Error! Reference source not found.). Going further, the highest rate of conversion from a completed smell test to a confirmed individual with hyposmia resulted from promoted online search results, earned press coverage, and radio ads (each with a conversion rate ≥20%; Error!

Reference source not found.). Those strategies had a greater likelihood of recruiting the target participant (i.e., those with hyposmia), but collectively only contributed 128 eligible participants with hyposmia. This may possibly be due to more limited reach of these approaches as compared to social media or other approaches that reach large general audiences. Key lessons learned thus far from PPMI recruitment are further detailed below.

First, it is valuable to remain agile with recruitment methods and adjust approaches in real time. Strategies may differ in success rates depending on the target population (i.e., those with a family history of PD vs. living with RBD). The level of study team time/resource investment for each approach must be considered in creating the appropriate mix of strategies for a study. The Foundation has iterated its approach with learnings on campaign return on investment. MJFF stopped investing in display and native ads (that appear beside or within text across websites) after analysis showed they drove page visits but did not translate into consents. In mid-2023, MJFF also pivoted away from resource-intensive engagement with senior living communities that introduced errors in research operations (e.g., assisted completion of consents). Future campaigns will utilize proven strategies (e.g., promoted social media posts), engage enriched populations (e.g., RBD individuals), and continue to test messaging and methods to enroll an inclusive study population.

Second, effective participant engagement at every step of the study process cannot be underestimated. Critical to enrollment for longitudinal follow-up — the study’s aim — was identification and continued engagement of hyposmic individuals, which is demonstrated by 42.6% (n=2,833) of all participants identified as hyposmic also agreeing to continue on to DAT imaging. We were successful in engaging participants to continue through the screening process to acquire DAT imaging through a combination of clear, comprehensive, and supportive communication, and assistance with travel planning for site visits, including longitudinal follow-up. Trained staff were provided talking points on the study details that helped retain hyposmic individuals for further screening with DaTscan. These talking points were centrally provided to ensure consistency and frequently iterated or expanded upon with learnings throughout the study.

Third, use of the centralized core to engage with the participants to provide structured systematic education, explanation of all results, and travel assistance was essential to ensure retention of the qualified individuals.

Fourth, it was evident that using centralized resources to reduce site burden was important and valuable to success. Site capacity was centrally monitored and adjusted as needed to redistribute work across sites to meet study needs for clinical baseline assessments, DaTscans, and subsequent clinical visits for longitudinal follow-up. In preparation for larger campaigns and anticipated spikes in demand, the study screening team was staffed to manage large volumes of participants through the centralized recruitment funnel. The centralized model allowed for a streamlined process to promptly shepherd participants through the screening workflow and ensure consistency in participants’ experiences throughout their journey. The model also allowed for timely reallocation of team resources during recruitment spikes to meet increased demands. Additionally, the flexibility of the centralized model allowed for rapid workflow implementations, including participant-facing content modifications in coordination with the MJFF communications team and relevant staff.

At this time, our recruitment efforts have helped meet ambitious study enrollment goals, and we hope they provide insights for other researchers. PPMI continues to recruit, and therefore we expect to continually iterate on our recruitment strategies. An important limitation of this approach is that PPMI benefits from a very large team and significant financial resources to achieve this scale of effort. It may not be possible to replicate this centralized approach to achieve broad recruitment for projects that are not as well resourced. As PPMI has deployed ST Direct in Europe and Canada, it is crucial to customize the operational activities to meet the needs of each setting/culture. We further recognize the significant limitations in our current methods to enroll a diverse study population and continue to develop and implement partnerships and strategies toward that measure. Finally, the majority of participants in ST Direct were female; strategies to encourage male participation would also need to be implemented.

We recognize the unique obstacles encountered in early disease studies of neurodegenerative disorders, including Alzheimer’s disease^13^ and PD^14–16^ given the fear and uncertainty associated with the diagnosis of a chronic and progressive neurological condition. Clear prerequisites to successful recruitment included targeted outreach to increase awareness of research opportunities in subgroups of interest coupled with strategic use of educational materials that promote health literacy and address the risk and benefits of study participation. The use of online and remote study protocols also holds great promise. We are hopeful that the insights from this extensive effort to recruit participants into PPMI described here will provide valuable perspective to other researchers encountering similar challenges enrolling participants at the very earliest stage of disease.

## Data Availability

Data used in the preparation of this article were obtained on March, 31, 2024 from the Parkinson’s Progression Markers Initiative (PPMI) database (www.ppmi-info.org/access-data-specimens/download-data), RRID:SCR_006431. For up-to-date information on the study, visit www.ppmi-info.org. This analysis was conducted by the PPMI Statistics Core and used actual dates of activity for participants, a restricted data element not available to public users of PPMI data.

## Acknowledgements

PPMI – a public-private partnership – is funded by the Michael J. Fox Foundation for Parkinson’s Research and funding partners, including 4D Pharma, Abbvie, AcureX, Allergan, Amathus Therapeutics, Aligning Science Across Parkinson’s, AskBio, Avid Radiopharmaceuticals, BIAL, BioArctic, Biogen, Biohaven, BioLegend, BlueRock Therapeutics, Bristol-Myers Squibb, Calico Labs, Capsida Biotherapeutics, Celgene, Cerevel Therapeutics, Coave Therapeutics, DaCapo Brainscience, Denali, Edmond J. Safra Foundation, Eli Lilly, Gain Therapeutics, GE HealthCare, Genentech, GSK, Golub Capital, Handl Therapeutics, Insitro, Jazz Pharmaceuticals, Johnson & Johnson Innovative Medicine, Lundbeck, Merck, Meso Scale Discovery, Mission Therapeutics, Neurocrine Biosciences, Neuron23, Neuropore, Pfizer, Piramal, Prevail Therapeutics, Roche, Sanofi, Servier, Sun Pharma Advanced Research Company, Takeda, Teva, UCB, Vanqua Bio, Verily, Voyager Therapeutics, the Weston Family Foundation and Yumanity Therapeutics. The authors sincerely appreciate the numerous and valuable contributions of the large number of study participants, as well as the study teams.

